# Dietary patterns of adults in Italy: Results from the third Italian National Food Consumption Survey, INRAN-SCAI

**DOI:** 10.1101/2024.10.18.24315728

**Authors:** Nicolò Scarsi, Roberta Pastorino, Cosimo Savoia, Gian Marco Raspolini, Angelo Maria Pezzullo, Stefania Boccia

## Abstract

**Background & objective.:** Diet is among the most significant modifiable risk factors for reducing the global burden of chronic diseases. This study aims to investigate the dietary patterns of adults in a large representative sample of the Italian population and to analyze these patterns according to sociodemographic characteristics.

**Methods:** Adult participants of the third Italian National Food Consumption Survey (INRAN- SCAI 2005-06) were included. A total of 878 food items were classified into 24 pre-defined food groups using the FoodEx2 classification system. Dietary patterns were identified through principal component analysis, and Z-scores were calculated to assess adherence to these patterns. Associations between sociodemographic characteristics, energy intake, and dietary adherence scores were investigated using linear regression models.

**Results:** Based on data from 2,831 subjects (median age 47, IQR 35-60), five principal components (PCs) were retained, explaining 35.6% of the overall variance. PC1 was indicative of a high-fat diet, PC2 suggested a western eating behavior, PC3 represented a health-conscious lifestyle, PC4 can be characterized as an Italian traditional diet, and PC5 represented an unhealthy dietary pattern. According to Z-scores, 42.4% of our study cohort showed high adherence to at least one of the dietary patterns. Less healthy dietary patterns were more prevalent among males and individuals from northern Italian regions.

**Conclusions:** Our results indicate a significant regional variation in terms of dietary pattern, mirroring the general trends of Italian eating habits of the last decades, characterized by a higher tendency towards a more westernized lifestyle. These findings underscore the importance of considering region-specific characteristics when designing future public health interventions and establishing national dietary guidelines.

## INTRODUCTION

Traditionally, nutritional epidemiology has investigated diet quality among individuals through analyses of micro- and macronutrients of single foods, which fail to consider the complexity of multiple food consumption (1). In contrast, multidimensional approaches, such as dietary pattern analysis, which allow the evaluation of the whole diet, have been a more effective way to reflect diet-health interconnections (2). A large body of literature reports on the role of certain poor dietary patterns and their potential threat to health (3–5). Several studies emphasized that a healthy diet, rich in fruits, vegetables, legumes, whole grains, and poor in free sugars, fats, especially saturated fats, and salt, can reduce the risk of several chronic diseases, as well as improve overall health and well-being (6–10). A systematic review and meta-analysis of randomized controlled trials found that a plant-based diet, which emphasizes fruits, vegetables, whole grains and legumes, and minimizes or excludes animal products, was associated with improvements in health outcomes (11). On the other hand, an unhealthy diet for adults (high in processed and refined foods, saturated and trans fats, added sugars and sodium) can increase the risk of chronic diseases and other health issues (12). Numerous studies thus far have highlighted the positive effects of the Mediterranean diet (13), which has been associated with numerous health benefits, including a reduced risk of cardiovascular disease, type 2 diabetes, and some cancers (14). However, recent data shows that Italian dietary habits have changed in recent years, and the adherence to the traditional Mediterranean diet has decreased. In this regard, according to a cross-sectional study in Italian adults, only about a fifth of the respondents reported high adherence rates to the Mediterranean diet, while the remaining part of the sample consumed a diet higher in saturated fats and added sugars. Furthermore, a recent systematic review, encompassing nine studies each featuring over 1,000 Italian participants, has demonstrated a consistent deviation from the Mediterranean diet over the past decade (15). The INRAN-SCAI survey was carried out from October 2005 to December 2006 on a representative sample of the Italian population and aimed to investigate the dietary habits of the Italian population, including the consumption of different food groups and nutrients (16) . The data collected in the context of such surveys can be informative on the dietary patterns of Italian populations, if properly analyzed (1,17). Empirical methods frequently used to derive dietary patterns include principal component analysis (PCA), reduced rank regression, and partial least squares regression analysis (17). PCA represents a well-known derivation method to investigate dietary patterns, which replaces a set of possibly correlating food groups with a new set of comprehensive indexes (principal components) that are uncorrelated and retain as much of the foods’ variance as possible (18). To our knowledge, while some studies investigated Italian dietary patterns (19–22), none so far used the INRAN- SCAI cohort, which represents the most updated available data source on individual food consumption in a nationally representative sample. Therefore, the aims of this study were (i) to derive main adult dietary patterns in the Italian population through PCA, using food frequencies data from the third Italian National Food Consumption Survey, INRAN-SCAI 2005-06, (ii) to characterize the dietary pattern adherence of the study sample using Z-scores and (iii) to explore the associations between these scores, sociodemographic characteristics, and energy intake.

## METHODS

### Study population and socio-demographic data

The third Italian National Food Consumption Survey, INRAN-SCAI 2005-06, is a cross-sectional survey based on a random sample of the general Italian population, previously described in detail elsewhere (16). Briefly, it involved a target sample of 1,329 households, with a 33% participation rate and aimed to characterize average food consumption in the four main Italy geographical areas (North-West, North-East, Center, and South and Islands). In total, food consumption data of 3,323 individuals and 9,984 daily food diaries, for 3 survey days, have been collected. INRAN-SCAI has been carried out by the National Research Institute on Food and Nutrition and was funded by the Italian Ministry of Agriculture, representing the third experience with food consumption surveys on a national scale. Apart from food consumption data, the following characteristics of the study sample were considered for the analysis: age, gender, body mass index (BMI, kg/m²), level of education and Italian area of residence. We considered only adult individuals (≥ 18 years) from INRAN-SCAI cohort participants. For the present analysis, we did not exclude participants based on implausible energy intake thresholds (e.g., <800 kcal/day) to retain the full sample of adults and capture a wider range of dietary patterns within the cohort.

### Food consumption data and food grouping

Food consumption was self-recorded by participants for three consecutive days using hard-copy diaries structured by meal, with all foods, beverages, food supplements, and ingested medicines registered. Individual food intakes were calculated using the software INRAN- DIARIO version 3.1. For each eating occasion, subjects were asked to record the time, place of consumption, detailed description of foods, quantity consumed, and brand, with portion sizes reported using a picture booklet for reference (16). Food frequency data from the third Italian National Food Consumption Survey, INRAN-SCAI 2005-06 were used to create a core food list containing 878 food items representing the diet of the sample under analysis. All food items have been classified into 24 pre-defined food groups according to a modified version of the food classification system FoodEx2. FoodEx2 contains descriptions of many food items aggregated into broader food groups and different levels of food categories (23). For the present analysis we excluded infant food (48 items) because only the adult population was considered, but it has been included either way in the database with single food items. In its original version, FoodEx2 is composed of 20 main food categories, but we modified it by classifying the ‘Meat’ items into six different subcategories, according to a meat classification suggested by Ferrari et al. (24). The extended list of the 24 pre-defined food groups, with some food items as example, is provided in Table 1.

**Table 1.** List of the 24 predefined food groups according to a modified version of the FoodEx2 Classification System.

| **Food groups** | **Examples of single items included** |
| --- | --- |
| Grain Products | Bread, crackers, pasta, rice, cereals, beignets, biscuits, flour, cake, croissant, couscous, processed wheat-based flakes |
| Vegetables | Garlic, puntarella, escaroles, carrots, onions, tomatoes, melons, turnips, mushrooms, aubergines, leeks, cardoons, lettuces, broccoli |
| Starchy Roots | Potatoes, potato flakes, potato starch |
| Legumes | Lentils, peas, peanuts, walnuts, coconuts, pistachios, pumpkin seeds |
| Fruits | Pineapples, avocado, kiwi, cherries, dried fruits, pears, peaches, jam of fruits, figs |
| Pork, Not Preserved, Excl. Offals | Wild boar meat, pig muscle, pig trotter, pig muscle, pig tissues, pig slaughtering products |
| Beef And Veal, Not Preserved, Excl. Offals | Cow fresh meat, bovine tongue, bull fresh meat, calf fresh meat |
| Offals, Blood, And Their Product | Brain, livers, edible offal of several flesh |
| Poultry And Game, Not Preserved, Excl. Offals | Goose, turkey, chicken, quail, pigeon, |
| Processed Meat | Meat in aspic, mortadella, ham, pancetta, cured seasoned meat |
| Other Meats, Not Preserved, Excl. Offal. | Goat, lamb, rabbit, horse, sheep, deer |
| Fish | Cod, squid, tuna, shrimps, sole, salmon, perch, trout, dentex, clam, scallops, canned fish, mullets |
| Dairy Products | Cheeses, yogurt, mozzarella, milk, ricotta, mascarpone, tofu |
| Egg Products | Eggs |
| Sugar, Confectionary | Honey, chocolate spread, candies, sucrose, toffee, chewing gum, nougat, bee-produced formulations |
| Fat And Oils | Oils, pork lard, butter, margarines |
| Juices | Fruit and vegetable juices, nectars without added sugars, food industry prepared |
| Non-Alcoholic Beverages | Cola beverages, infusions, tea, soft drinks, multivitamin juices |
| Alcoholic Beverages | Beer, wine, liqueurs, brandy, vermouth, Marsala |
| Water | Natural mineral water, carbonated bottled drinking water, tap water |
| Herbs, Spices, Condiment | Capers buds, parsley, ginger roots, celery leaves, vinegar |
| Special Nutritional | Mixed supplements vitamins, herbal formulations, proteins, dietary foods for special medical purposes |
| Composite Food | Sandwich bread, soups, mixed vegetable salad, pizza, and pizza like dishes |
| Snacks Etc | Wafers, custard, desserts, popcorn, ice cream, sorbet, potato crisps |

### Statistical analyses

All analyses were performed using STATA software, v.16.1 (Stata Corp., College Station, Texas, USA, 2019). The categorical variables are presented by absolute and relative frequencies (n and %). Numerical variables were described by median and interquartile range (IQR). Data distribution was assessed by visual inspection and using the Shapiro-Wilk test. PCA was performed to derive dietary patterns (17). We used as input variables log-transformed residuals obtained from linear regression models (Residual Method), in which the explanatory variable was the median kilocalories intake, and the response variable was the median energy intake of each participant (25,26). This approach has been used to control the role of energy intake. The number of principal components was selected based on Kaiser criterion (Eigenvalue > 1) and on the scree-plot criterion as an ancillary method (27). Every dietary pattern has a factor loading for each food group, and we employed Varimax rotation and Kaiser normalization to factor loadings to facilitate data interpretability (28). Large positive or negative factor loadings suggest the foods that are important in that component; positive loadings greater than 0.2 and negative loadings lower than 0.2 were considered in the interpretation of principal components to characterize and label dietary patterns (18). To evaluate individual diet adherence to the dietary patterns identified, Z-scores ((individual value - mean)/SD) were calculated. In depth, the individual values were determined by summing up observed intakes of each food group, weighted by the respective factor loading. Subsequently, we have described median food intake (g/day) of individuals who showed a dietary adherence equal to or greater than 1 standard deviation above the average of the study sample (Z-Score ≥ 1) for each identified dietary pattern. To investigate the determinants of higher (or lower) dietary pattern Z-scores, linear regression models were firstly applied for demographic variables alone (unadjusted analysis). Subsequently, a comprehensive analysis was conducted using linear regression models, where Z-scores served as the dependent variable. The independent variables included demographic factors such as age, sex, education level, geographical area and average daily energy intake. Both unadjusted and adjusted regression coefficients were calculated and reported with their respective 95% confidence intervals (CI). In the adjusted analysis, we included average daily energy intake as an additional independent variable to control for potential bias due to the relationship between higher food consumption and greater diet adherence, which could influence Z-scores. For the present analysis, we decided not to adjust for covariates or potential confounders. This decision was made to focus on the direct relationships between the dependent variable (Z-scores) and the independent variables (e.g., demographic factors, energy intake), acknowledging that unadjusted models may not account for confounding effects.

## RESULTS

### Participant characteristics

Sociodemographic characteristics of the sample are shown in Table 2. Out of the 3,323 individuals involved in the INRAN-SCAI cohort, 492 (18.8%) participants, all of them aged under 18, were excluded from the analysis. The final study sample consisted of 2,831 individuals, of which 1,561 (55.1%) were female. The median age of the sample was 47 years (IQR 35-60). The participants were from South and Islands (n=996; 35.2%), North-West (n=738; 26.1%), North-East (n=559; 19.7%) and Center regions (n=538; 19.0%). Most of our population (n=1,683; 63.1%) had a secondary education, followed by those who had a tertiary (n=552; 20.7) and primary (n=431; 16.2%) education level. Over half of adults (n=1,657; 58.5%) showed a healthy BMI (18.5-24.99 kg/m²). Concerning the eating habits of the study population, the median energy intake was 1900.2 kcal (IQR: 1572.4 – 2286.9 kcal), and the median carbohydrate intake was 250.1g/d (IQR: 198.5 – 304.2g/d). In contrast, the median intakes of total fat, saturated fatty acids (SFA) and fiber were 82.3g/d (IQR: 66 – 99.7 g/d), 25.1g/d (IQR: 19.3 – 31.6 g/d) and 17.9g/d (IQR: 14.2 – 22.3 g/d), respectively”

**Table 2.** Baseline characteristics of the study sample.

|  | **Total (N = 2831)** |
| --- | --- |
| Age, median (IQR) | 47 (35 - 60) |
| Energy intake (kcal), median (IQR) | 1900.2 (1572.4 - 2286.9) |
| BMI (kg/m<sup>2</sup>), median (IQR) | 24.1 (22 - 26.6) |
| Carbohydrates (g/d), median (IQR) | 250.1 (198.5 - 304.2) |
| Fiber (g/d), median (IQR) | 17.9 (14.2 - 22.3) |
| Total fat (g/d), median (IQR) | 82.3 (66 - 99.7) |
| SFA (g/d), median (IQR) | 25.1 (19.3 - 31.6) |
| **Gender, n (%)** |  |
| Male | 1270 (44.9) |
| Female | 1561 (55.1) |
| **BMI (kg/m<sup>2</sup>), n (%)** |  |
| Underweight | 80 (2.8) |
| Normal | 1657 (58.5) |
| Overweight | 856 (30.2) |
| Obese | 238 (8.5) |
| *Missing* | 1 (0.03) |
| **Level of education, n (%)** |  |
| Primary | 431 (16.2) |
| Secondary | 1683 (63.1) |
| Tertiary | 552 (20.7) |
| *Missing* | 165 (5.8) |
| **Geographical area, n (%)** |  |
| North-West | 738 (26.1) |
| North-East | 559 (19.7) |
| Center | 538 (19.0) |
| South & islands | 996 (35.2) |

\*1 missing value for BMI
\*\*165 missing values for education level

### Dietary patterns

According to the PCA results, five principal components (PC) were retained, explaining the 35.6% of the overall variance. The KMO measure verified the sampling adequacy for the analysis (KMO = 0.71), indicating that the correlation was adequate for PCA. PC1 was reminiscent of a high-fat diet characterized by positive loadings for vegetables, starchy roots, egg products, fats and oils, and low amounts of alcoholic beverages. PC2 depicted a “Western” eating behavior, positively associated with processed meat, offals, other meats, juices, special nutritional products, composite food, and snacks. PC3 represented a health-conscious diet, positively loaded by vegetables, fruits and water, negatively associated with pork meat and alcoholic beverages consumption. PC4 had similarities with an Italian-like diet, positively loaded by grain products, vegetables, herbs, spices, and condiments. PC5 represented an unhealthy dietary pattern, positively associated with sugar and confectionery, non-alcoholic beverages and negatively correlated with vegetable consumption. Main characteristics of the five dietary patterns, and related food groups and factor loadings are reported in Table 3 and in supplementary material (S1), respectively.

**Table 3.** Characteristics of the 5 dietary patterns derived from the PCA on the quantitative variables of 24 food groups (g/day).

| Principal component (PC) | Factor loadings | Cumulated explained variance | PC label |
| --- | --- | --- | --- |
| PC1 | Vegetables, starchy roots, beef and veal, eggs products, fat and oils, alcoholic beverages | 9.27% | High-fat |
| PC2 | Processed meat, offal, other meats, juices, special nutritional, composite food, snack etc. | 17.55% | Western |
| PC3 | Vegetables, fruits, water (pork, alcoholic beverages) * | 23.72% | Health-conscious |
| PC4 | Grain products, vegetables, herbs, spices, and condiments | 29.80% | Italian, traditional |
| PC5 | Sugar and confectionary, non-alcoholic beverages (vegetables)* | 35.63% | Junk, out of meal |

*\*Negative loadings are specified between parentheses*

### Dietary pattern Z-scores

According to Z-scores, 1,202 (42.4%) adults of our study cohort showed high adherence to at least one of the dietary patterns, of which 644 (53.6%) were males and 458 (38%) found to be overweight or obese. Median daily food intakes (g/day) of individuals with Z-Score ≥ 1, according to dietary patterns and to the 24 predefined food groups, are reported in Table 4. Out of those that adhere the most (Z-Score ≥ 1), 410 (34.1%), 371 (30.8%), 369 (30.7%), 426 (35.4%), 396 (32,9%) individuals were attributed to the PC1, PC2, PC3, PC4 and PC5, respectively. Additional information is reported in the Supplementary material (S2).

**Table 4.**
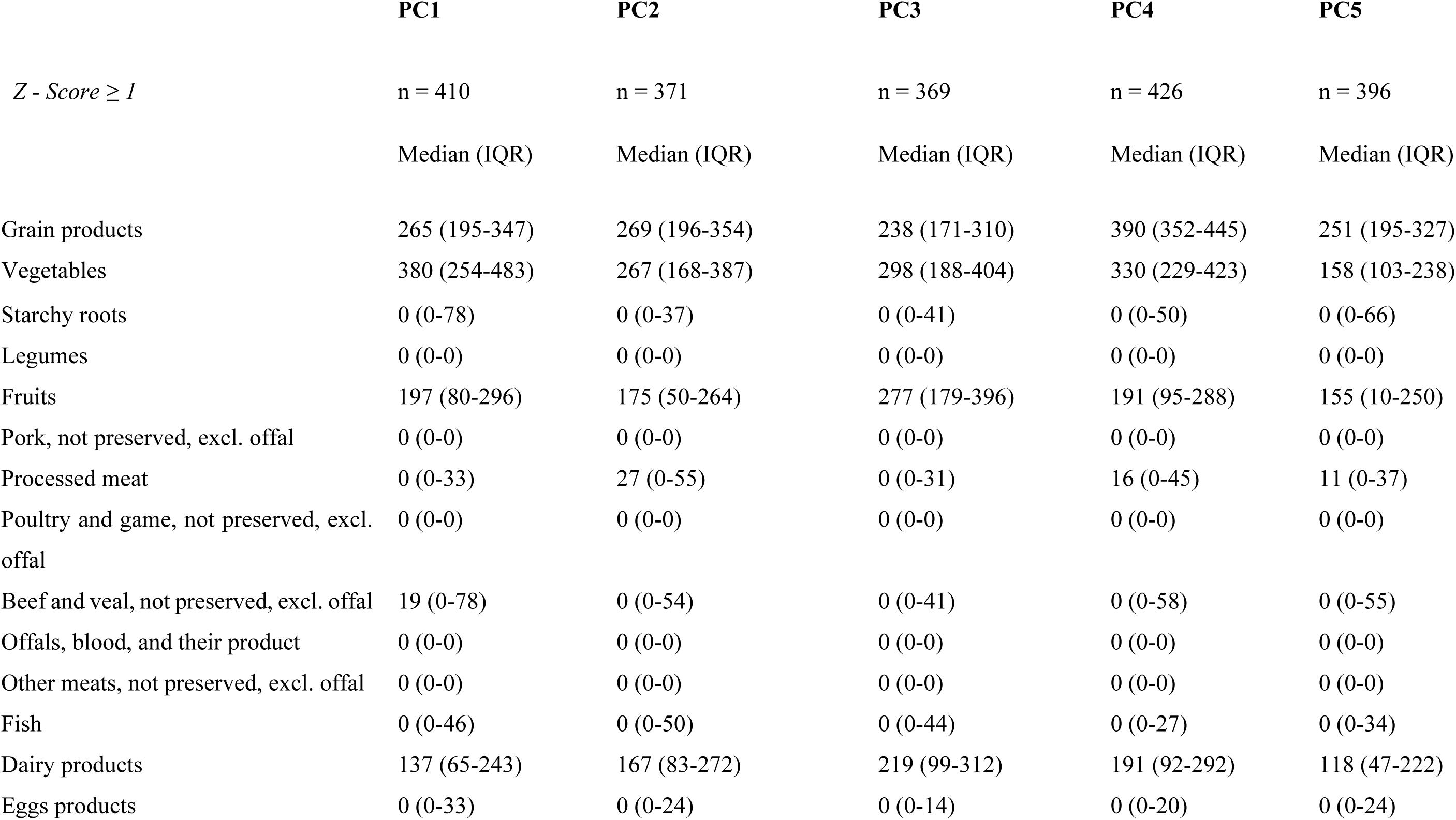
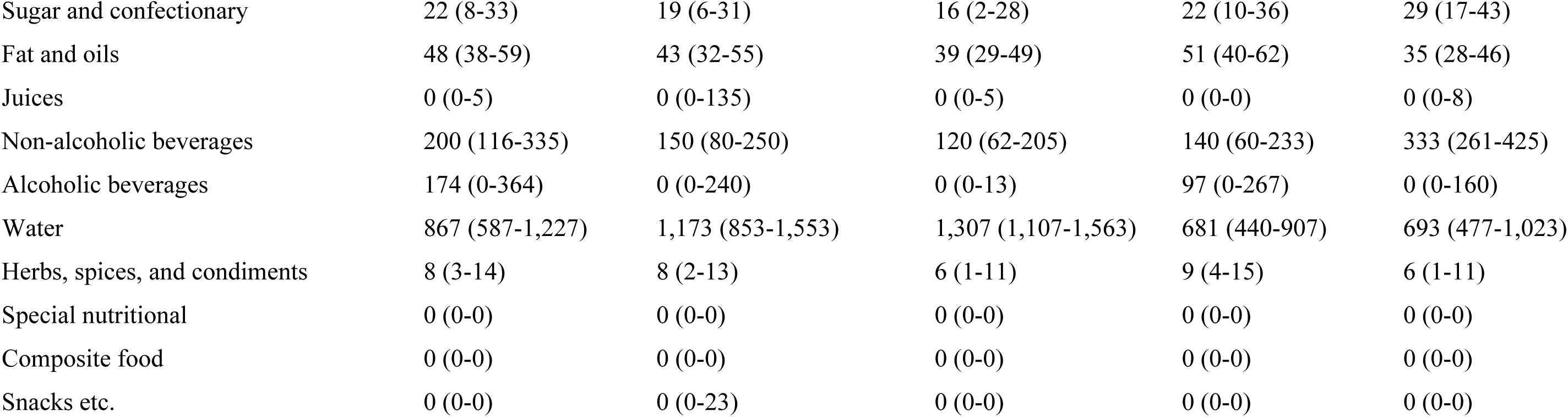
Food intake (g/day) of individuals with high adherence (Z - Score ≥ 1) to dietary patterns according to the 24 predefined food groups.

|  | **PC1** | **PC2** | **PC3** | **PC4** | **PC5** |
| --- | --- | --- | --- | --- | --- |
| *Z* - Score $\geq 1$ | n = 410 | n = 371 | n = 369 | n = 426 | n = 396 |
|  | Median (IQR) | Median (IQR) | Median (IQR) | Median (IQR) | Median (IQR) |
| Grain products | 265 (195-347) | 269 (196-354) | 238 (171-310) | 390 (352-445) | 251 (195-327) |
| Vegetables | 380 (254-483) | 267 (168-387) | 298 (188-404) | 330 (229-423) | 158 (103-238) |
| Starchy roots | 0 (0-78) | 0 (0-37) | 0 (0-41) | 0 (0-50) | 0 (0-66) |
| Legumes | 0 (0-0) | 0 (0-0) | 0 (0-0) | 0 (0-0) | 0 (0-0) |
| Fruits | 197 (80-296) | 175 (50-264) | 277 (179-396) | 191 (95-288) | 155 (10-250) |
| Pork, not preserved, excl. offal | 0 (0-0) | 0 (0-0) | 0 (0-0) | 0 (0-0) | 0 (0-0) |
| Processed meat | 0 (0-33) | 27 (0-55) | 0 (0-31) | 16 (0-45) | 11 (0-37) |
| Poultry and game, not preserved, excl. offal | 0 (0-0) | 0 (0-0) | 0 (0-0) | 0 (0-0) | 0 (0-0) |
| Beef and veal, not preserved, excl. offal | 19 (0-78) | 0 (0-54) | 0 (0-41) | 0 (0-58) | 0 (0-55) |
| Offals, blood, and their product | 0 (0-0) | 0 (0-0) | 0 (0-0) | 0 (0-0) | 0 (0-0) |
| Other meats, not preserved, excl. offal | 0 (0-0) | 0 (0-0) | 0 (0-0) | 0 (0-0) | 0 (0-0) |
| Fish | 0 (0-46) | 0 (0-50) | 0 (0-44) | 0 (0-27) | 0 (0-34) |
| Dairy products | 137 (65-243) | 167 (83-272) | 219 (99-312) | 191 (92-292) | 118 (47-222) |
| Eggs products | 0 (0-33) | 0 (0-24) | 0 (0-14) | 0 (0-20) | 0 (0-24) |
| Sugar and confectionary | 22 (8-33) | 19 (6-31) | 16 (2-28) | 22 (10-36) | 29 (17-43) |
| Fat and oils | 48 (38-59) | 43 (32-55) | 39 (29-49) | 51 (40-62) | 35 (28-46) |
| Juices | 0 (0-5) | 0 (0-135) | 0 (0-5) | 0 (0-0) | 0 (0-8) |
| Non-alcoholic beverages | 200 (116-335) | 150 (80-250) | 120 (62-205) | 140 (60-233) | 333 (261-425) |
| Alcoholic beverages | 174 (0-364) | 0 (0-240) | 0 (0-13) | 97 (0-267) | 0 (0-160) |
| Water | 867 (587-1,227) | 1,173 (853-1,553) | 1,307 (1,107-1,563) | 681 (440-907) | 693 (477-1,023) |
| Herbs, spices, and condiments | 8 (3-14) | 8 (2-13) | 6 (1-11) | 9 (4-15) | 6 (1-11) |
| Special nutritional | 0 (0-0) | 0 (0-0) | 0 (0-0) | 0 (0-0) | 0 (0-0) |
| Composite food | 0 (0-0) | 0 (0-0) | 0 (0-0) | 0 (0-0) | 0 (0-0) |
| Snacks etc. | 0 (0-0) | 0 (0-23) | 0 (0-0) | 0 (0-0) | 0 (0-0) |

### General linear models

Regression model coefficients and 95% confidence intervals for socio-demographic characteristics, along with energy intake, are provided in Table 5 (adjusted and unadjusted). The text that follows summarizes the key findings.

**Table 5.**
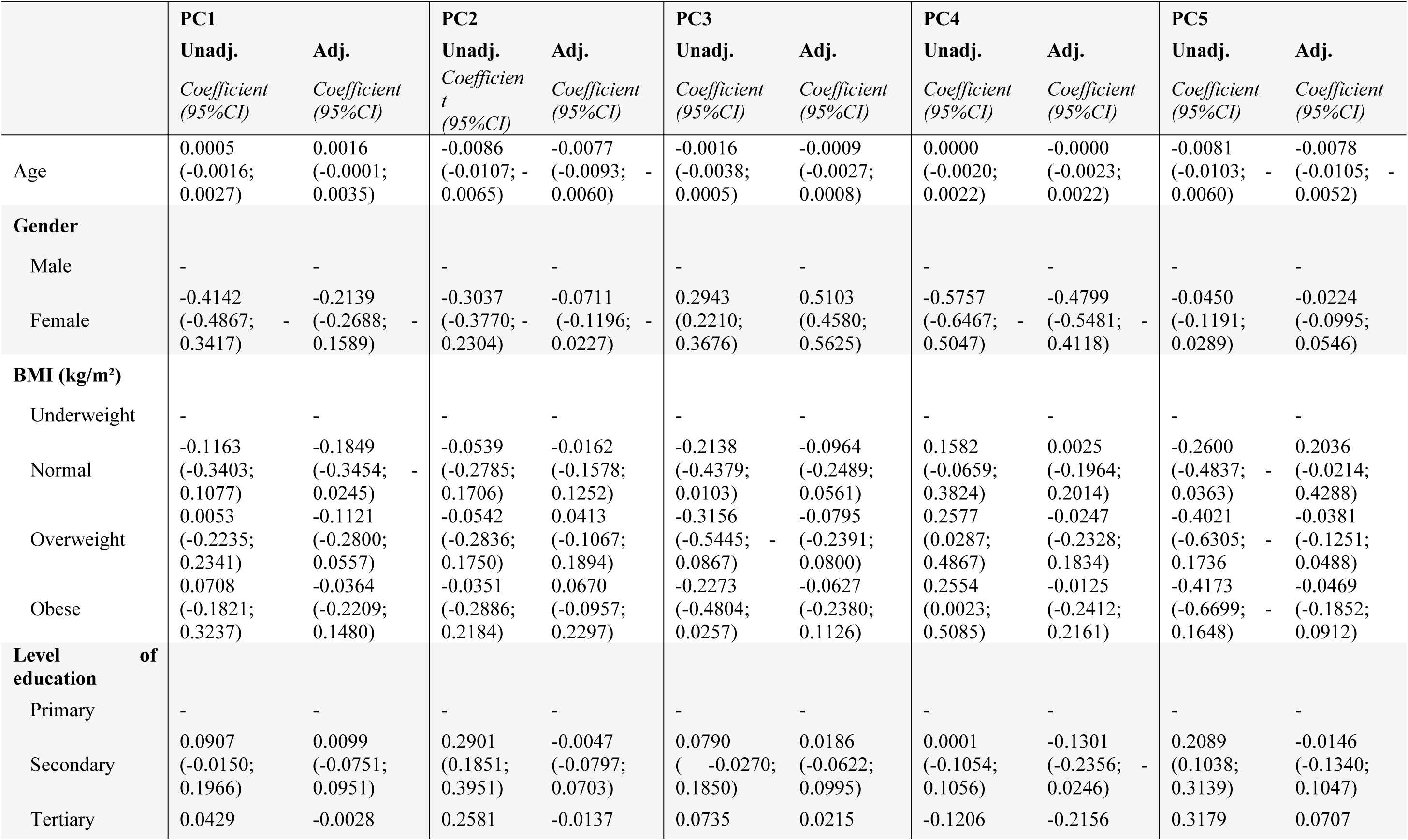
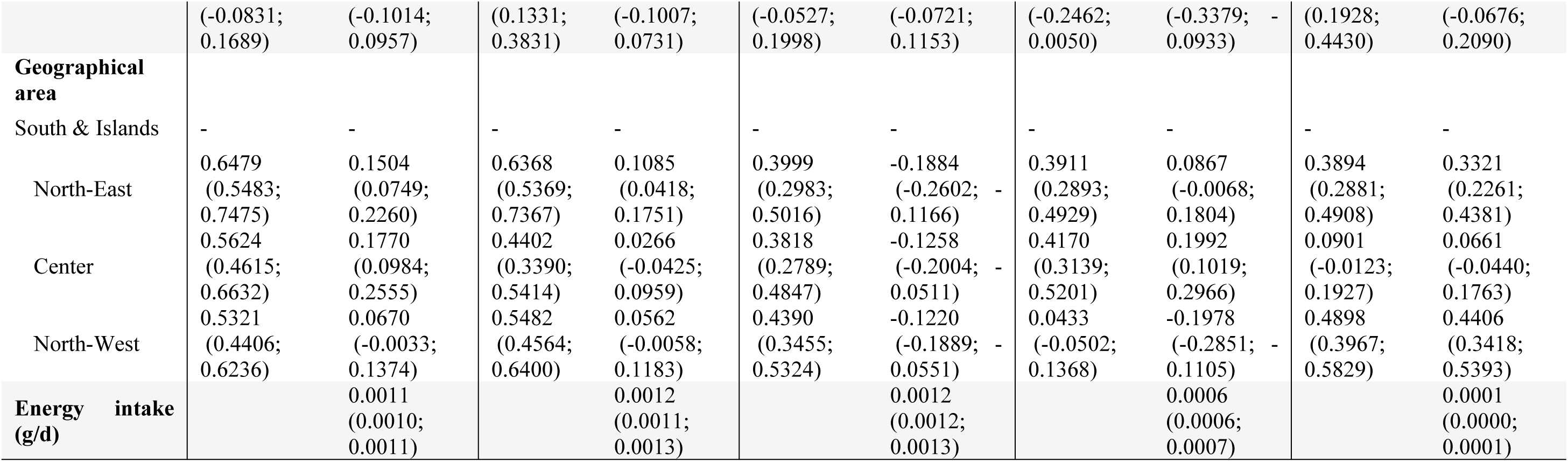
Regression model coefficients and 95% confidence intervals for socio-demographic characteristics.

### Pattern 1 - High-fat diet

Coming from the North-East (β = 0.1504, p < 0.001) or Center regions (β = 0.1770, p < 0.001) was associated with higher adherence to the ’High fat’ dietary pattern compared to being from the South & Islands. Conversely, being female (β= -0.2139; p<0.001) and normal weight (β= - 0.1849; p=0.024) were associated with lower dietary adherence scores.

### Pattern 2 - Western

Coming from the North-East (β= 0.1085; p=0.001) was associated with higher adherence to the “Western” dietary pattern compared to being from the South & Islands. Conversely, age (β= -0.0077; p<0.001) and female gender (β= -0.0711; p= 0.004) were associated with lower dietary adherence scores.

### Pattern 3 - Health conscious

Female gender (β= 0.5103; p<0.001) was associated with higher Z-scores on the “Health conscious” dietary pattern. Inversely, coming from North-West (β= -0.1220; p<0.001), North- East (β= -0.1884; p<0.001) or center regions (β= -0.1258; p=0.001) were associated with lower dietary adherence scores compared to being from the South & Islands.

### Pattern 4 - Italian traditional

Coming from center regions (β= 0.1992; p<0.001) was associated with higher Z-scores on the “Italian traditional” dietary pattern compared to being from the South & Islands. On the other hand, being female (β= -0.4799; p<0.001), having either a secondary (β= -0.1301; p=0.016) or tertiary (β= -0.2156; p= 0.001) education level and coming from North-West regions (β= - 0.1978; p<0.001) were associated with lower dietary adherence scores.

### Pattern 5 - Junk, out of meal

Coming from North-East (β= 0.3321; p<0.001) and North-West (β= 0.4406; p<0.001) regions were associated with higher Z-scores on the “Junk, out of meal” dietary pattern compared to being from the South & Islands. Age (β= -0.0078; p<0.001) was associated with lower dietary adherence scores.

## DISCUSSION

This study aimed to investigate adult dietary patterns in Italy within the INRAN-SCAI cohort and to understand the characteristics of individuals adhering closely to these patterns. Our analysis revealed a diverse range of dietary habits among adults, identifying five major patterns: “High-Fat,” “Western,” “Health-Conscious,” “Traditional,” and “Junk, Out of Meal.” These patterns collectively accounted for approximately one-third of the variance in food intake, with around 42.4% of participants demonstrating high adherence to at least one identified pattern. Factors such as gender, age, BMI, education level, and regional location explained differences in dietary behavior, with less healthy dietary patterns being more prevalent among males and individuals from northern Italian regions. The dietary patterns identified in this study are consistent with those previously reported over the past two decades in both adult and adolescent Italian populations (29). Specifically, the “Western” and “Junk, Out of Meal” patterns share characteristics with a major dietary pattern known as “Western style”, which is characterized by a high intake of foods rich in saturated fats, added sugars, and processed foods, typical of the Western diet. On the other hand, the “Health-Conscious,” “High-Fat,” and “Italian, Traditional” patterns resemble the “Mediterranean-style” diet historically representative of Mediterranean countries, characterized by a high consumption of fruits, vegetables, whole grains, fish, legumes, and olive oil. However, it’s important to note a degree of overlap among dietary patterns, with individuals often exhibiting high adherence to multiple patterns simultaneously. Previous research by Edefonti et al. provided evidence of similar variability in dietary patterns across European cohorts, suggesting the robustness of dietary pattern analysis in different populations in terms of reproducibility and stability over time (30). In our analysis, individuals from northern regions displayed less healthful dietary choices compared to those in central or southern regions, highlighting potential interregional differences influenced by factors such as crop diversity, socio-economic status, and urbanization. Similar regional dietary variations have been observed in other European countries (31–33). For instance, studies in Croatia have documented differences in dietary patterns between coastal areas and urban centers, while Switzerland has shown variations in food consumption across German-, French-, and Italian-speaking cantons. These observations underscore the importance of considering regional factors in public health interventions promoting healthy eating habits. Additionally, our study identified associations between certain demographic factors and dietary patterns. Overall, being female was associated with higher Z-scores on the “Health conscious” dietary pattern, and with lower adherence scores on the unhealthier dietary patterns, respectively. Older individuals were more likely to exhibit a “High Fat” diet, aligning with global nutrition surveys reporting higher intakes of saturated fats and cholesterol in older adults (34). Conversely, individuals with tertiary education were associated with a “Junk, Out of Meal” pattern, a finding that warrants further investigation given the lack of scientific evidence on the higher intake of dietary sugars among this demographic group.

### Strengths and limitations

The present study has some strengths and limitations that warrant acknowledgment and may influence the interpretation of the results. Firstly, the relatively short duration of the survey, covering only a 3-day period, may lead to an overestimation of long-term food consumption. Another limitation arises from the decision to not adjust for covariates or potential confounders. While this approach allowed us to focus on the direct relationships between Z-scores and the independent variables, it may not fully account for potential confounding effects, which could influence the observed associations. Other limitations include subjective decisions related to data-driven methods, such as the determination of the number of principal components to retain and dietary pattern labeling. Such decisions can significantly influence the comparability of results across studies, as the characteristics of dietary patterns identified may vary based on labeling criteria and methodological choices related to input variables quantification, format, transformation, food grouping schemes, and rotation of factor loadings (36).

## CONCLUSIONS

This study represents the first attempt to identify dietary patterns using Principal Component Analysis with data from the third Italian National Food Consumption Survey INRAN-SCAI 2005-06 cohort. Two out of five (“Western” and “Junk, Out of Meal”) distinct dietary patterns emerged, with evidence suggesting a prevalence of unhealthy dietary behaviors, particularly among males. Notably, adults from northern Italian regions exhibited a preference for less healthy dietary patterns compared to those in central or southern regions, indicative of a lifestyle characterized by increased consumption of meat, snacks, and sugar-sweetened beverages. These findings underscore the importance of considering region-specific characteristics when designing future public health interventions and establishing national dietary guidelines. Integrating insights from dietary pattern analysis studies into policy recommendations is crucial for promoting healthier eating habits and reducing diet-related health risks within the Italian population.

## Declaration of competing interest

The authors declare that there is no conflict of interest.

## Authors’ Contributions

NS, AMP, CS, GMR, RP, SB contributed to the establishment of the present work. NS, AMP downloaded INRAN-SCAI 2005-06 data from the Global Dietary Database. AMP, GMR, CS supported NS in writing the manuscript. NS made data preparation and analysis. AMP, RB, SB, critically revised the article, approved the version to be published and agreed to be accountable for all aspects of the work ensuring integrity and accuracy.

## Data Availability

All relevant data are within the manuscript and its Supporting Information files.

## ACKNOWLEDGMENTS

The authors are most thankful to the Italian Ministry of Agriculture who funded the INRAN- SCAI 2005–06 survey, to the Italian households who participated in this study and to all field workers who made this work possible.

## Funding

This paper is part of the PhD Project of Nicolò Scarsi, under the PON “Research and Innovation” 2014-2020. ACTION IV.5 “Doctorates on Green Issues: “*Development of A Tool To assess Adherence to The Planetary Health Diet Proposed by The Eat-Lancet Commission*”. Funder had no role in the establishment of the present study.

**Correspondence and requests for materials** should be addressed to Roberta Pastorino

**Additional information** Supplementary materials

- S1) Supplementary Table 1. Estimated factor loadings from PCA.
- S2) Table showing adherence to multiple dietary patterns simultaneously.

## Notes

### Competing Interest Statement

The authors have declared no competing interest.

### Funding Statement

Yes

### Author Declarations

This study was conducted according to the guidelines laid down in the Declaration of Helsinki and all procedures involving research study participants were approved by the ethics committee. Written informed consent was obtained from all subjects/patients.

